# The Potential Impact of Intensified Community Hand Hygiene Interventions on Respiratory tract Infections: A Modelling Study

**DOI:** 10.1101/2020.05.26.20113464

**Authors:** Thi Mui Pham, Yin Mo, Ben S. Cooper

## Abstract

Increased hand hygiene amongst the general public has been widely promoted as one of the most important non-pharmaceutical interventions for reducing transmission during the ongoing COVID-19 pandemic and is likely to continue to play a key role in long-term efforts to suppress transmission before a vaccine can be deployed. For other respiratory tract infections community hand hygiene interventions are supported by evidence from randomised trials, but information on how effectiveness in reducing transmission scales with achieved changes in hand hygiene behaviour is lacking. This information is of critical importance when considering the potential value of substantially enhancing community hand hygiene frequency to help suppress COVID-19. Here, we developed a simple model-based framework for understanding the key determinants of the effectiveness of changes in hand hygiene behaviour in reducing transmission and use it to explore the potential impact of interventions aimed at achieving large-scale population-wide changes in hand hygiene behaviour. Our analyses show that the effect of hand hygiene is highly dependent on the duration of viral persistence on hands and that hand washing needs to be performed very frequently or immediately after hand contamination events in order to substantially reduce the probability of infection. Hand washing at a lower frequency, such as every 30 minutes or with a delay of 15 minutes after contamination events, may be adequate to reduce the probability of infection when viral survival on hands is longer, such as when hands are contaminated with mucus. Immediate hand washing after contamination is more effective than hand washing at fixed-time intervals even when the total number of hand washing events is similar. This event-prompted hand washing strategy is consistently more effective than fixed-time strategy regardless of hand contamination rates and should be highlighted in hand hygiene campaigns.

## Introduction

Promotion of hand hygiene is a key public health intervention in preventing the spread of infectious diseases. Since the mid-1800s, when Ignaz Philip Semmelweis demonstrated that hand washing could dramatically reduce maternal mortality due to puerperal fever [1], hand hygiene has been the cornerstone of infection prevention and control policies. In the hospital setting, hand hygiene has played a major role in successfully controlling hospital-acquired infections, especially those caused by methicillin-resistant *Staphylococcus aureus* [2]. In the community, there is evidence from randomised controlled trials that hand hygiene interventions can be effective in reducing both the risk of diarrhoeal disease [3] and respiratory infections [4, 5, 6].

In response to the coronavirus disease 2019 (COVID-19) pandemic, hand hygiene has been re-emphasized as a primary focus in public information campaigns [7, 8, 9]. Hand hygiene is simple, low-cost, minimally disruptive and, when widely adopted, may lead to substantial population-level effects [5, 10]. While randomised controlled trials of hand hygiene interventions in the community provide evidence that such interventions are effective in reducing the incidence of respiratory tract infections, reported effect sizes are highly variable [4, 6]. It is unclear to what extent this variability is explained by success in achieving substantial changes in hand hygiene behaviour in these trials. Understanding how the effectiveness of hand hygiene in reducing transmission scales with hand hygiene frequency is important for assessing the potential contribution to COVID-19 suppression efforts of interventions that aim to achieve a large and sustained increase in community hand hygiene.

At present, most public health agencies recommend washing hands “more often” than prior to the pandemic, and after coughing or sneezing, but specific indications of precisely how frequently to wash hands are generally lacking (see Table 1). This lack of more specific recommendations reflects the lack of a quantitative understanding of how different levels of hand hygiene behaviour affect transmission risk.

**Table 1.** Hand hygiene recommendations advised by major public health agencies.

| Organisation | Public health message |  | Ref |
| --- | --- | --- | --- |
|  | General public | Households with suspected/ confirmed patients |  |
| WHO | Regularly and thoroughly clean hands with alcohol-based hand rub or soap and water, especially after coughing or sneezing, when caring for the sick, before, during and after food preparation, before eating, after toilet use, when hands are visibly dirty. | Family members: Wash hands with soap and water regularly, especially after coughing or sneezing, before, during and after food preparation, before eating, after toilet use, before and after caring for ill persons and when hands are visibly dirty.<br>Patients: Wash hands immediately and thoroughly after coughing, sneezing, removing face mask. Stay in a separate room from other family members. | [11] |
| ECDC | Rigorous hand-washing with soap and water >20 seconds, or alcohol-based solutions, gels or tissues is recommended in all community settings in all possible scenarios, especially after coughing or sneezing, disposal of used tissues. | Family members: Wash hands frequently, especially after contact with the patient or with any surface frequently touched by the patient, e.g., before and after preparing food, before eating, after using the toilet, removing face mask/ gloves, handling waste.<br>Patients: Wash hands immediately and thoroughly after coughing, sneezing, removing face mask. | [12, 13] |
| PHE | Washing hands more often, especially after arriving at work or home, after blowing nose, coughing or sneezing, before eating or handling food. | Wash hands frequently with soap and water for 20 seconds or using hand sanitiser, especially after coughing/sneezing and disposal of used tissue. | [14, 15] |
| CDC | Wash hands often with soap and water for >20 seconds especially after being in a public place, or after blowing nose, coughing, or sneezing. | Clean frequently touched surfaces and objects daily (e.g., tables, countertops, light switches, doorknobs, and cabinet handles) using a regular household detergent and water. Wash hands often with soap and water for >20 seconds or hand sanitizer, especially after going to the bathroom, before eating, and after blowing nose, coughing, or sneezing. Always wash your hands with soap and water if your hands are visibly dirty. | [16, 17] |

Abbreviations: WHO - World Health Organization; ECDC - European Centre for Disease Prevention and Control; PHE - Public Health England (UK); CDC - Centers for Disease Control and Prevention (US)

In this study, we take a theory-based approach and develop a simple mechanistic mathematical model to understand the relationships between the various components of respiratory tract infection transmission pathways involving hand contamination. Finally, we consider the implications of the outcomes of these analyses for the potential contribution of intensified community hand hygiene to the suppression of severe acute respiratory syndrome coronavirus 2 (SARS-CoV-2) and, more generally, for reducing respiratory tract infections with other pathogens.

## Methods

### Overview

We consider infections that are mediated by contaminated hands and initially neglect direct droplet or aerosol transmission. Hands are assumed to become contaminated with infectious material via contact with contaminated surfaces or an infected person. In the absence of hand washing, hands do not remain contaminated indefinitely; instead, as has been shown experimentally, the probability of remaining contaminated and capable of transmitting infection declines over time (Figure 1, top panel) [18, 19]. If contaminated hands of a susceptible host make contact with the host’s mucous membranes in the eyes, nose or mouth there is some probability of the host becoming infected. Effective hand washing interrupts this process by removing viable pathogens from the hands.

**Figure 1.**
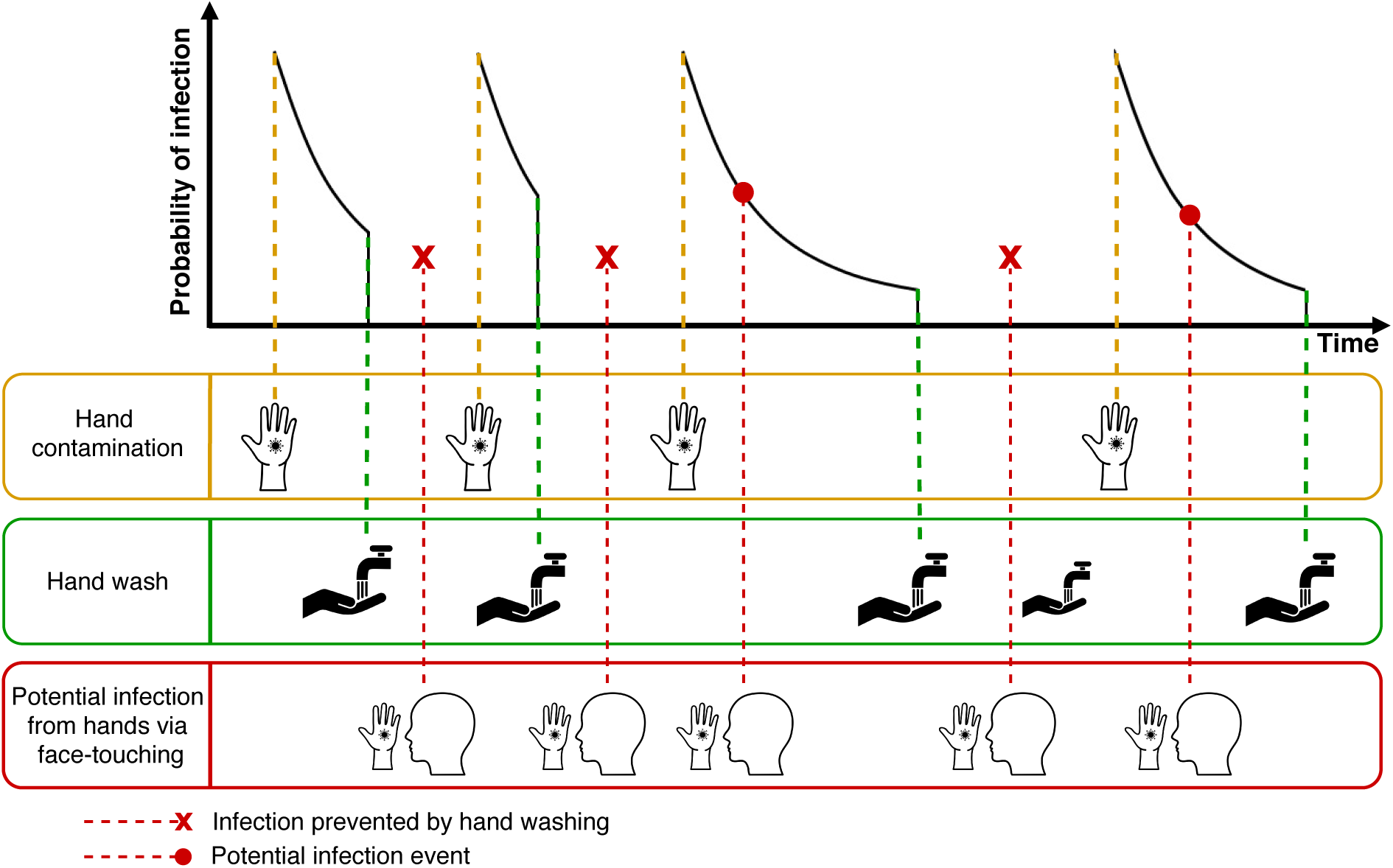
Hand hygiene model. Illustration of potential infection events from hands via face-touching, hand contamination events, and hand washing events. Hand contamination events increase the probability of infection, which then decreases exponentially with time. Hand washing eliminates the probability of infection during subsequent face-touching if no further hand contamination events occur. An infection may occur between a hand contamination event and hand washing, depending on the probability of infection at the time of face-touching.

An immediate consequence of this conceptualisation is that the time interval between the hands becoming contaminated and making infectious contact with the host’s mucosa can have a critical impact on how effective a given frequency of hand washing will be at interrupting transmission (Figure 2). If this time interval is relatively long in the absence of hand hygiene, regular effective hand hygiene will have a high chance of blocking potential transmission events (red diamonds in Figure 2 panel A). In contrast, if this time interval is short much more frequent hand hygiene will be needed to block an appreciable proportion of transmission events (Figure 2 panel B). Given a certain probability of infection, the time interval between hand contamination and transmission to the host’s mucosa tends to be longer if virus persistence on hands is long and vice versa.

**Figure 2.**
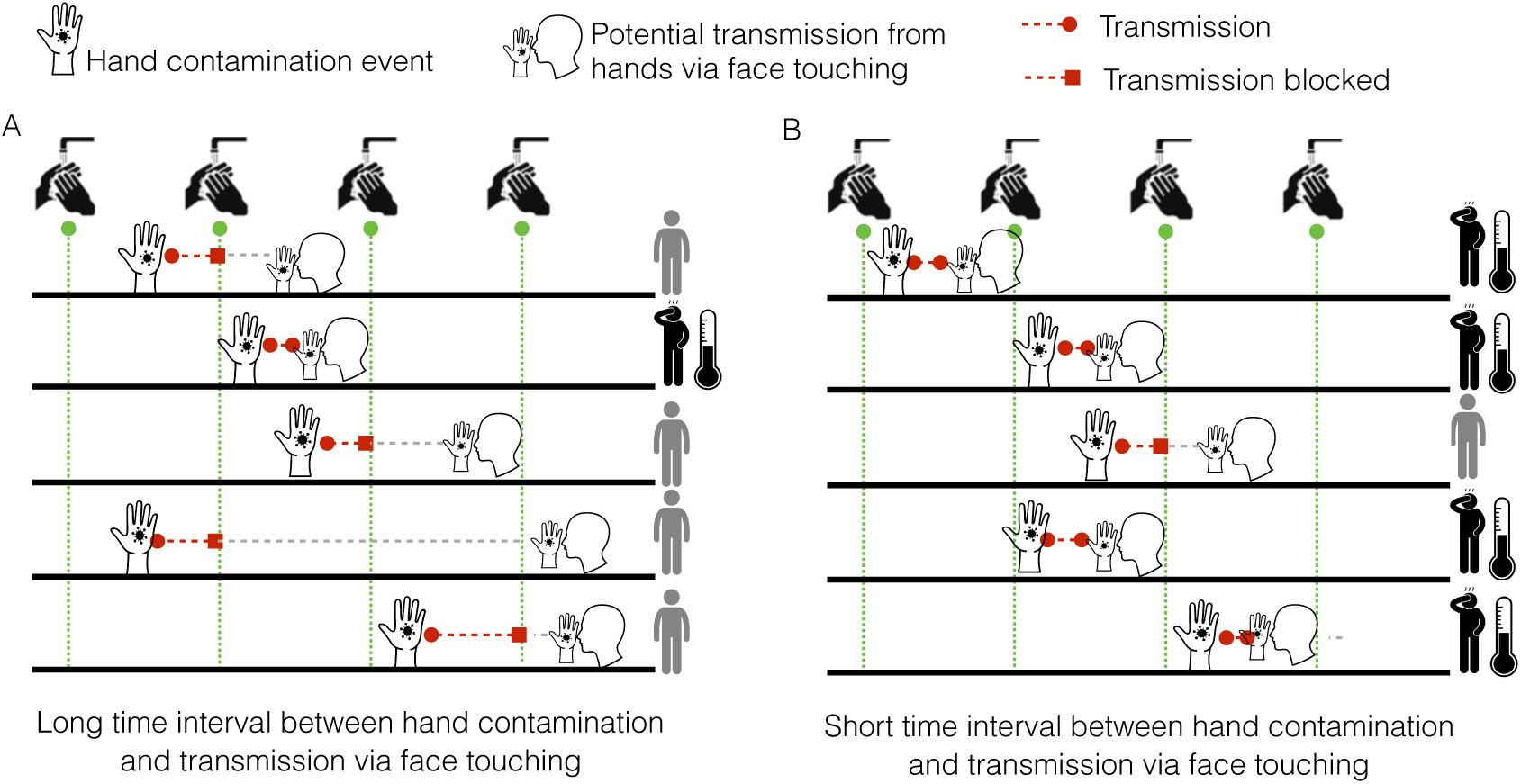
Long versus short time interval between hand contamination and infection with regular hand washing. A) When there are long time intervals between hand contamination and potential infection from hands via face-touching, hand washing can block many infection events and substantially reduce the risk of infection. B) When there are short time intervals between hand contamination and face-touching, it is likely that hand washing can disrupt only a few infections.

### Hand hygiene scenarios

We varied the timing of hand washing in the following schemes:

1. uniformly at fixed time intervals (fixed-time hand washing), or
2. with a delay after hand contamination events (event-prompted hand washing).

### Mathematical model

Hands of susceptible individuals are assumed to become contaminated at random. These contamination events are assumed to occur independently of each other, and to follow a Poisson distribution with a mean of λ*_c_* events per hour. The probability of the virus persisting on hands at time *t* after contamination, *P*(*t*), is assumed to decay exponentially with a half-life of *T*_1/2._ This is consistent with experimental data for influenza A (see [19]). Individuals touch their face at random leading to potential infection events that are assumed to occur independently of each other, and follow a Poisson distribution with a mean of *λ_f_* events per hour. The probability that a single face-touching contact with contaminated hands actually leads to transmission is *ε*.

Assume the face-touching events occur at times *t*_1_,… *,t_F_* during the given time period *T*. Then the cumulative probability of infection over the time period *T* is given by:

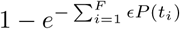

We assume that when hand washing is performed after the last hand contamination event and before a face-touching event at time *t_i_*, the respective probability of virus persistence *P*(*t_i_*) is reduced to zero. We make the conservative assumption that hand washing by infectious individuals does not alter their chance of infecting others since infectious individuals may transmit the virus to others without direct physical contact. A detailed mathematical description of the model is included in the supplementary material.

### Parameters

When available, parameter estimates were obtained from the literature. Otherwise, we performed sensitivity analyses where parameters were varied within plausible ranges (see Table 2). In the fixed-time hand washing scheme, we varied time intervals between hand washing to be 5 min to 6 hours. For event-prompted hand washing, the delay of hand washing after hand contamination events was varied from 1 min to 6 hours.

**Table 2.**
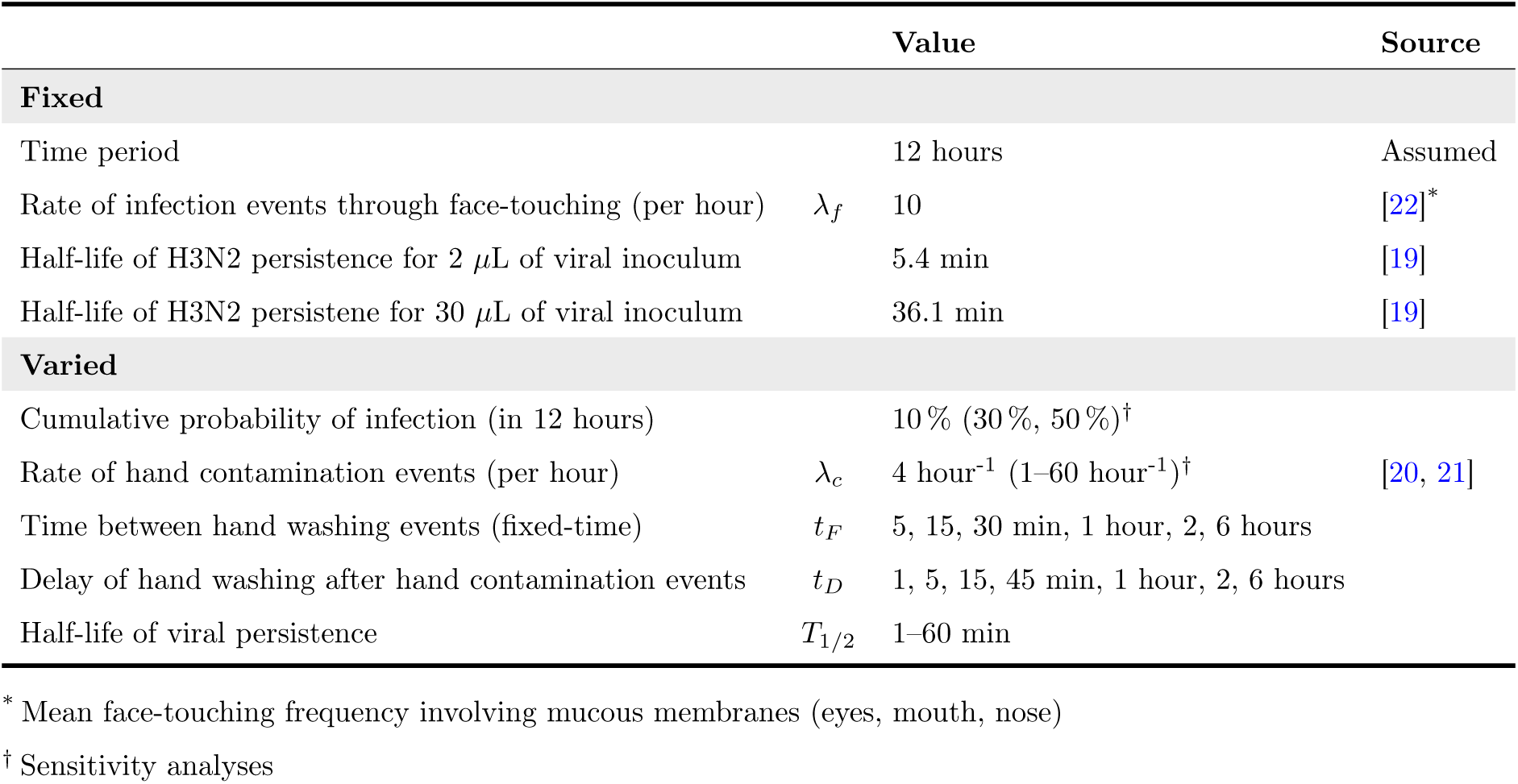
Parameter values.

|  |  | Value | Source |
| --- | --- | --- | --- |
| **Fixed** |  |  |  |
| Time period |  | 12 hours | Assumed |
| Rate of infection events through face-touching (per hour) | $\lambda_f$ | 10 | [22]* |
| Half-life of H3N2 persistence for 2 μL of viral inoculum | | 5.4 min | [19] |
| Half-life of H3N2 persistence for 30 μL of viral inoculum | | 36.1 min | [19] |
| **Varied** |  |  |  |
| Cumulative probability of infection (in 12 hours) |  | 10 % (30 %, 50 %)† |  |
| Rate of hand contamination events (per hour) | $\lambda_c$ | 4 hour<sup>-1</sup> (1–60 hour<sup>-1</sup>)† | [20, 21] |
| Time between hand washing events (fixed-time) | $t_F$ | 5, 15, 30 min, 1 hour, 2, 6 hours | |
| Delay of hand washing after hand contamination events | $t_D$ | 1, 5, 15, 45 min, 1 hour, 2, 6 hours | |
| Half-life of viral persistence | $T_{1/2}$ | 1–60 min | |

\* Mean face-touching frequency involving mucous membranes (eyes, mouth, nose)
† Sensitivity analyses

There is little published data on the rate of hand contamination events susceptible individuals are exposed to when in contact with infected individuals who are shedding respiratory viruses, and none specific to SARS-CoV-2. In a direct observation study conducted by Zhang et al [20], surface touching behaviour in a graduate student office was recorded. Approximately 112 surface touches per hour were registered. Another study by Boone et al [21] found that the influenza virus was detected on 53% of commonly touched surfaces in homes with infected children. Informed by these values, we took 60 events per hour as the upper bound for the rate of hand contamination events *λ_c_*. We chose 1 event per hour as the lower bound. In our main analyses, we used a rate of 4 hand contamination events per hour.

To date, it is not known how long SARS-CoV-2 can persist on human fingers. In [19], the survival of influenza A on human fingers was experimentally investigated. We fitted exponential decay curves to these results in order to determine the half-life of probability of persistence of H3N2 for two viral volumes of 2 *μ*L and 30 *μ*L (see Table 2 and supplementary material). In addition, we vary the half-life of probability of persistence from 1 to 60 min in our analysis.

### Model outcomes

The model output is the cumulative probability of a susceptible person becoming infected in twelve hours and we will refer to it subsequently as simply the probability of infection. We investigated the impact of hand washing on the probability of infection for different hand contamination rates. In addition, we compared the two hand washing schemes (fixed-time vs. event-prompted) to find the optimal hand washing strategy that will lead to the greatest reduction of the probability of infection. The model was implemented in R version 3.6.3 [23]. The code reproducing the results of this study is available at https://github.com/tm-pham/covid-19_handhygiene.

## Results

Viral persistence on hands plays a key role on the effect of increasing hand hygiene frequency. The longer the virus survives on the hands, the larger the impact of increasing hand washing uptake on the probability of infection. For example, when the half-life of viral persistence is 1 min, hand washing every 15 min reduces the probability of infection from 10 % to 9.2 % (Figure 3 Panel A). When the half-lives increase to 5.4 min and 36.1 min (equivalent to the half-lives of H3N2 persistence of 2 *μ*L and 30 *μ*L viral inoculum, respectively), the same hand washing frequency decreases the probability of infection to 6.9% and to 4.6 %, respectively. Consequently, fewer hand washes are necessary to reduce the probability of infection by 50 % for long compared to short durations of viral persistence (see Figure S2). This observation can be explained by the fact that the shorter the virus persists on hands, the shorter the intervals between hand contamination and transmission events tend to be (with a higher transmission probability per contact, see Figure S4) and therefore, the less likely hand washing is able to interrupt infection events. Figure S3 shows that the total time of exposure between hand contamination and hand washes increases with increasing half-life of viral persistence, confirming the hypothesis that timely hand washing is especially crucial if the virus survives only a short time on fingers. Furthermore, the effect of hand washing on reducing the probability of infection plateaus with increasing duration of virus persistence (Figure 3). This can be attributed to the hand contamination rate, i.e. new events occur before the virus decays.

**Figure 3.**
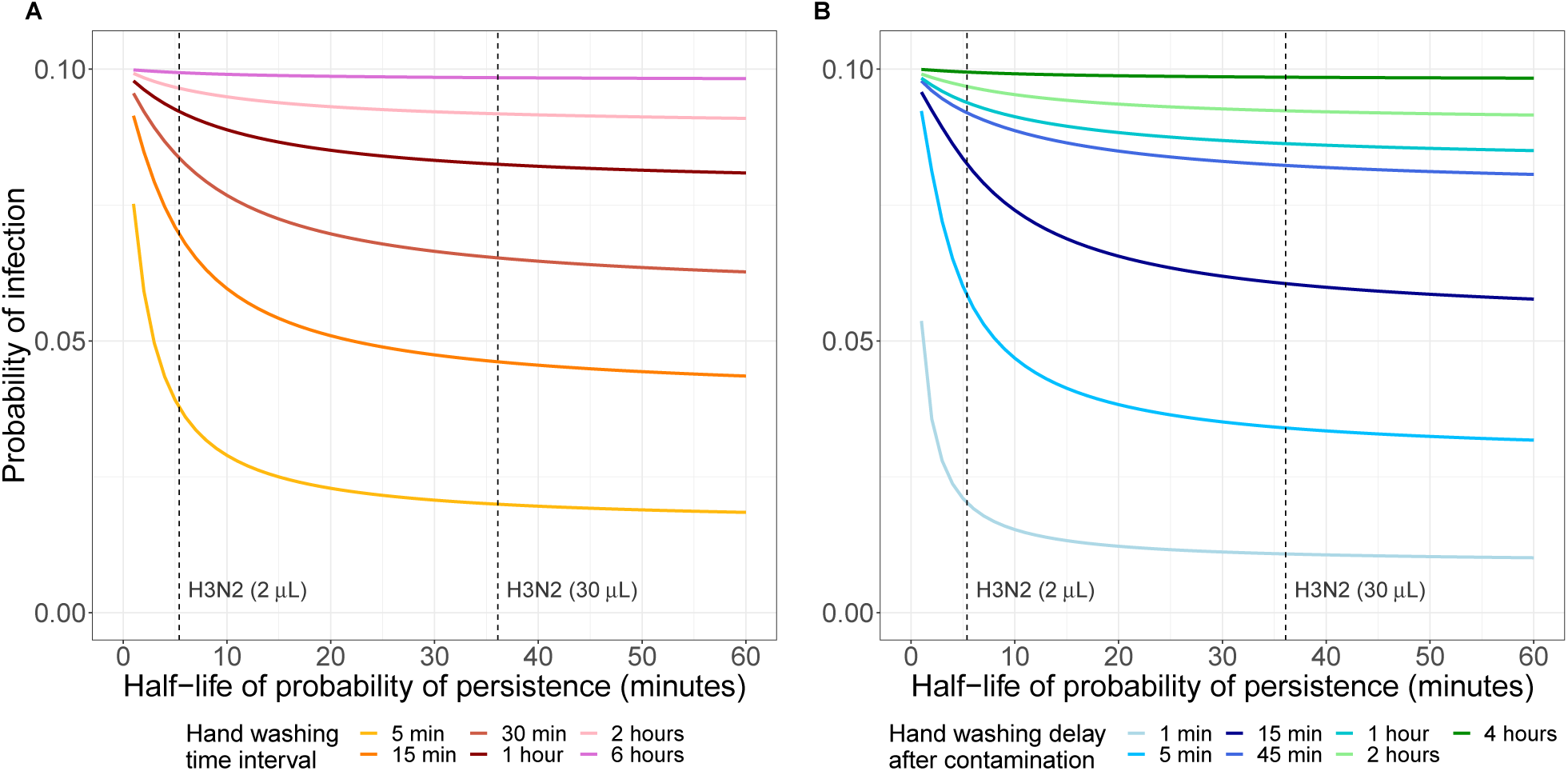
Impact of half-life of viral persistence on probability of infection for different hand washing schemes and frequencies. (A) Fixed-time hand washing (B) Event-prompted hand washing. In this graph, we assumed that a susceptible individual is exposed to a baseline probability of infection of 10% if no hand washing is performed within the time period of twelve hours. The dashed lines represent the half-life of viral persistence for H3N2 inoculum volumes of 2 *μ*L and 30 *μ*L (calculated from [19]). For each half-life value, the probability of transmission per face-touching event *ε* was determined for a probability of infection = 10% in the case of no hand washing. The probability of infection for the different hand washing frequencies/delays was then computed using this *ε* value. Hand contamination events are assumed to occur on average 4 times per hour. Sensitivity analyses with different values for baseline probabilities of infection as well as the half-life calculations are presented in the supplementary material.

The second notable finding from the model is that event-prompted hand washing is more effective than fixed-time hand washing in reducing the probability of infection. We illustrate this in Figure 4 by comparing both schemes using four different hand washing frequencies/delays, each with approximately the same average number of hand washing performed per hour. For example, hand washing regularly every fifteen minutes is compared to event-prompted hand washing one minute after each hand contamination event (set at four per hour). If the half-life of viral persistence is similar to 2 *μ*L of H3N2 inoculum (*T*_1/2_ = 5.4 min), the baseline probability of infection of 10%(no hand washing) is reduced to about 6% and 2% when hand washing is performed every 15min and one minute after hand contamination events, respectively. The differences between the two hand washing schemes are less pronounced if hand washing is performed less frequently or with a longer delay after hand contamination events since the two hand washing schemes become more similar. It follows that delays between hand contamination and hand washing decrease the effect of hand washing on reducing the probability of infection.

**Figure 4.**
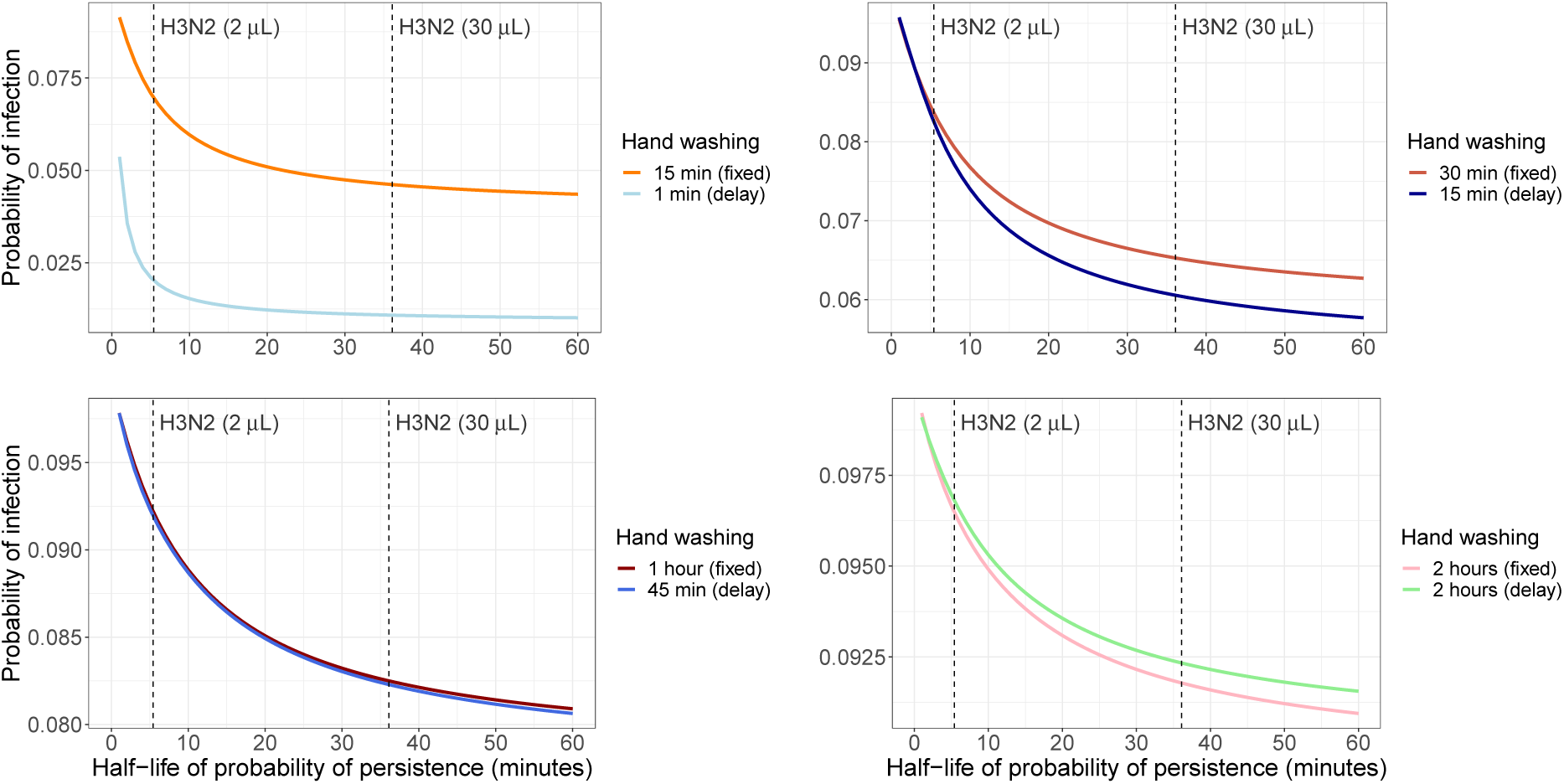
Comparison of the impact of the two hand washing schemes on the cumulative probability of infection. Hand washing at fixed time intervals and event-prompted hand washing with similar average number of hand washing events per hour are compared for a hand contamination rate of *λ_c_* =4 hour^-1^. A baseline probability of infection of 10% is assumed when there is no hand washing. The dashed lines represent the half-life values of H3N2 persistence for 2 *μ*L and 30 *μ*L inoculum volumes [19].

Another important parameter that affects the effect of hand hygiene is the hand contamination rate. Figure 5 shows the increase in hand hygiene frequency required to half the probability of infection from 10% (no hand wash) to 5%. When hand contamination rate is relatively rare, at less than 10 times per hour, fewer hand washes are needed to reduce the probability of infection if hand washing is performed event-prompted. In addition, the longer the virus persists on hands, the smaller the number of hand washes have to be performed to reduce the probability of infection. This effect is less pronounced for event-prompted than for time-fixed hand washing, re-emphasizing the finding that when hand contamination occurs very frequently, hand washing would need to be very frequent to have an impact on reducing the probability of infection, e.g., at least five times per hour to prevent 50% of transmission (in the case of a half-life of 36.1 min). In this case, susceptible individuals are exposed to a continuous risk of hand contamination and hand washing has only a limited impact on reducing the risk of infection.

**Figure 5.**
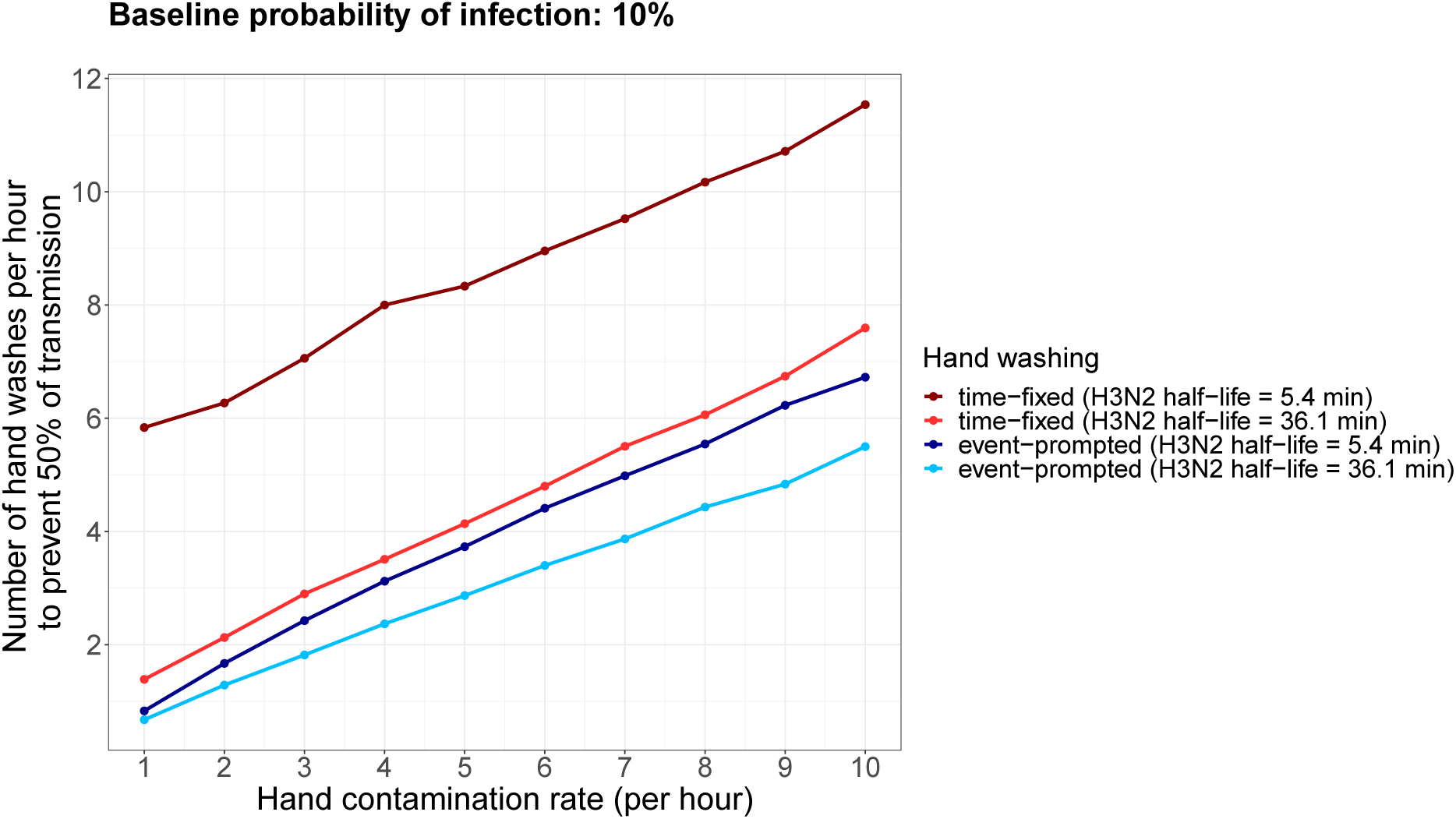
Number of hand washes necessary to prevent 50% of transmission. For a baseline probability of infection of 10%, the number of hand washing events necessary to reduce the probability of infection to 5% was computed for time-fixed and event-prompted hand washing and a range of hand contamination rates. We used the half-life of H3N2 persistence for viral inoculum volumes of 2 *μ*L and 30 *μ*L (calculated from [19]).

We performed sensitivity analyses for different baseline probabilities of infection and hand contamination rates (see supplementary material). Our qualitative conclusions do not change with respect to different baseline probabilities of infection.

## Discussion

Our study provides new insights into factors that affect the effectiveness of hand hygiene behaviour in reducing the probability of infection. Firstly, we found that the shorter the virus survives on hands, the less effective increasing hand hygiene frequency is in reducing infection. The logic behind this is that when the virus dies off quickly before hand washing is performed, the time intervals between hand contamination and transmission tend to be shorter and the respective transmission probability per contact tends to be higher for the same cumulative probability of infection. Secondly, contaminated surfaces are crucial for the effect of hand hygiene. The more often hands become contaminated, the more frequently hands need to be washed to reduce infection risk. Lastly, when hands are not constantly contaminated, event-prompted hand washing is more efficient than fixed-time hand washing given the same hand washing frequency. This is because delays in hand washing after contamination of hands in fixed-time compared to event-prompted hand washing tend to be longer, and, during this delay, susceptible hosts may become infected through face-touching.

These findings provide additional insights behind the modest and heterogeneous effects of hand hygiene reported by hand hygiene trials aimed at reducing respiratory tract infections in the community [4, 6, 24]. These trials are challenging to conduct due to the difficulties in implementing behaviour change, including poor adherence to hand washing recommendations [25], loss-to-follow up [26, 27], and external factors influencing behaviour such as the 2009 H1N1 outbreak [28].

Because the hand contamination rate directly impacts the effect of hand hygiene, specific hand hygiene advice should cater for different situations where surface contamination differs markedly. For example, contacts in the community and in a household with an infectious person would be likely to result in very different hand contamination rates. In the first case, where hand contamination events occur at a moderate rate, hand washing needs to be performed frequently or immediately after hand contamination events in order to substantially reduce the probability of infection. To facilitate this, hand sanitisers should ideally be installed or provided in public areas with high-touch surface areas, such as public transportation and supermarkets, to reduce the delay in hand cleansing. Although this has been recommended by the World Health Organization during the ongoing COVID-19 pandemic [29], many national governments have not prioritized the easy access to hand hygiene facilities. Furthermore, in the second case, where hands become contaminated very frequently, a substantial reduction in the probability of infection is unlikely to be attained unless hand washing frequency is increased drastically, i.e., every one to five minutes. Because hand washing at such a high rate is not practical, the recommendation in this scenario is to regularly clean the environment and/or isolate infected individuals to reduce hand contamination events.

Our model has several limitations. Firstly, we specifically modelled indirect transmission routes via hands and did not consider direct droplet and aerosol transmission. To date, there is little known about the relative importance of the various transmission routes of respiratory pathogens [30]. When other routes are considered, the effect of hand hygiene will be reduced. Secondly, there is limited literature on many parameters used in the model, which prevents us from making more precise quantitative conclusions. These include the probability of infection with contaminated hands, the survival of pathogens on contaminated hands and infective dose. Furthermore, we modelled all infection events with the same rate of decay, i.e., the same probability of pathogen persistence on the hands. In reality, hand contamination events are likely to be heterogeneous with small droplets persisting only a short amount of time and heavy contamination with mucus decaying at a slower rate. We performed sensitivity analyses with varying parameter values and distributions to ensure our conclusions are robust on a qualitative level.

## Data Availability

The code reproducing the results in the manuscript is available here.

https://github.com/tm-pham/handhygiene_modelling

## Supplementary material

### Model

Hands of susceptible individuals are assumed to get contaminated at random. These contamination events are assumed to occur independently of each other, and follow a Poisson distribution with a mean of *λ_c_* events per hour. The probability of the virus to persist on hands at time *t* after contamination, *P*(*t*), is assumed to decay exponentially with a half-life of *T*_1/2_. This is consistent with experimental data for influenza A (see [19]). Individuals touch their face at random leading to potential infection events that are assumed to occur independently of each other, and follow a Poisson distribution with a mean of *λ_f_* events per hour. The probability that a single face-touching contact with contaminated hands actually leads to transmission is *ε*.

Assume the face-touching events occur at times *t*_1_,…, *t_F_* during the given time period *T*. Then the cumulative probability of infection over the time period *T* is given by:

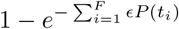

We assume that when hand washing is performed after the last hand contamination event and before a face-touching event at time *t_i_*, the respective probability of virus persistence *P*(*t_i_*) is reduced to zero.

#### Probability of viral persistence on contaminated hands

The decay of the probability of viral persistence on contaminated hands is modeled as an exponential decay with probability distribution:

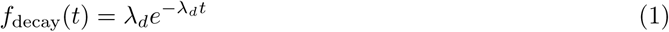

where *λ_d_* is the decay constant. The probability that virus will die off within time *t* is given by the integral of the decay distribution function from 0 to *t*:

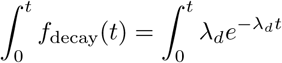

The probability that the virus will persist at time *t* is one minus the probability that it will die off within the same period:

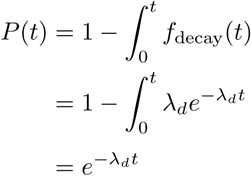

The average survival time (or mean lifetime) is given by:

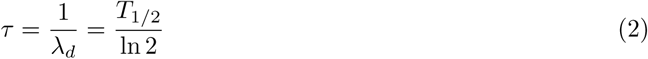

## Estimation of virus half-life

We estimated the half-life of viral survival on contaminated hands using experiments conducted by Thomas et al,[19] where 2 *μ*L and 30 *μ*L of influenza A (H3N2) viral suspension mixed with respiratory secretions were deposited on finger tips. The half-lives were calculated using data from both the 2 *μ*L ([19] Figure 2) and 30*μ*L ([19] Figure 3) H3N2 viral inoculum experiments with an exponential decay model:

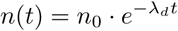

where *λ_d_* is the decay rate. The decaying quantity, *n*(*t*), represents the number of fingers with recoverable infectious viral particles and is assumed to have an initial value of *n*_0_ at time zero.

In the experiment with 2 *μ*L inoculum, 18 contaminated fingers from six individuals were tested for the presence of infectious virus at 1, 3, 5, 15 and 30min after initial contamination. Figure S1 depicts the data and the fitted curve for the 2 *μ*L inoculum. The decay rate was estimated to be 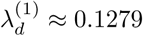. The half-life is therefore given by 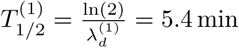.

For 30 *μ*L of viral inoculum, a total of 12 fingers were contaminated and the presence of H3N2 was tested after 15 min. We estimated the half-life by using these two data points (see Figure 3 in [19]). Thus, 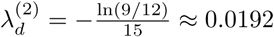. Therefore, 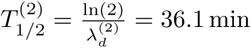.

**Figure S1.**
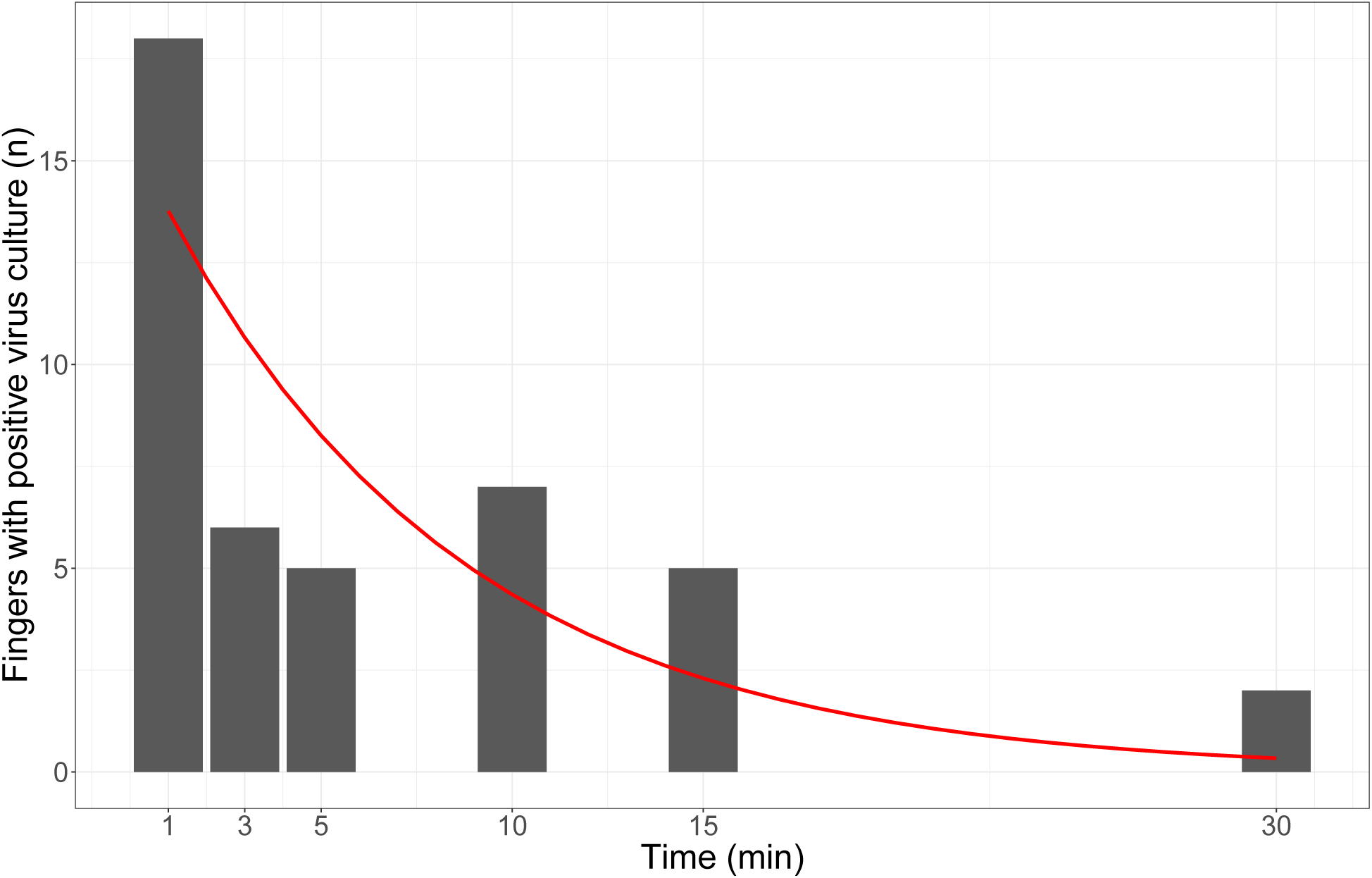
Influenza A(H3N2) virus survival on fingers over time. Data was retrieved from [19]. A 2 *μ*L drop of influenza A (H3N2) viral suspension mixed with respiratory secretions was deposited on fingertips. Bars represent the absolute number of fingers from which infectious virus was recovered. The red line represents the exponential decay curve *n*(*t*) = 15.65*e*^-0.1279^*^t^* fitted to this data.

## Hand washing and half-life of virus persistence

The shorter the half-life of virus persistence, the higher the frequency of hand washing necessary in order to prevent 50% of infections (see Figure S2). In addition, the time intervals between hand contamination and hand washes have to be shorter in order to prevent 50% of the infections (see Figure S3.

**Figure S2.**
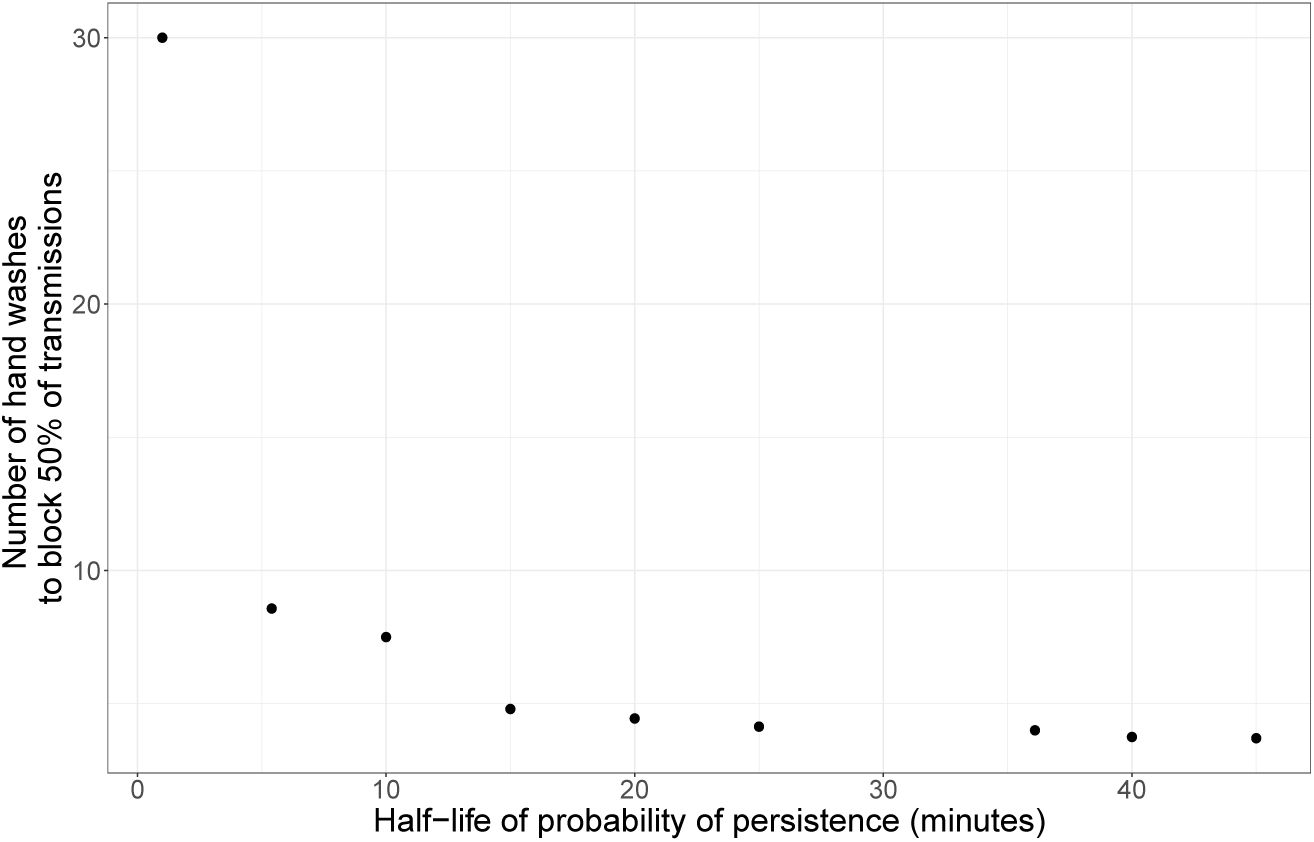
Number of fixed-time hand washes necessary to prevent 50% of transmissions. For each half-life value of virus persistence, the number of hand washes that is necessary to prevent 50% of transmission was computed for a baseline probability of infection of 10%. Hand contamination events are assumed to occur on average 4 times per hour.

**Figure S3.**
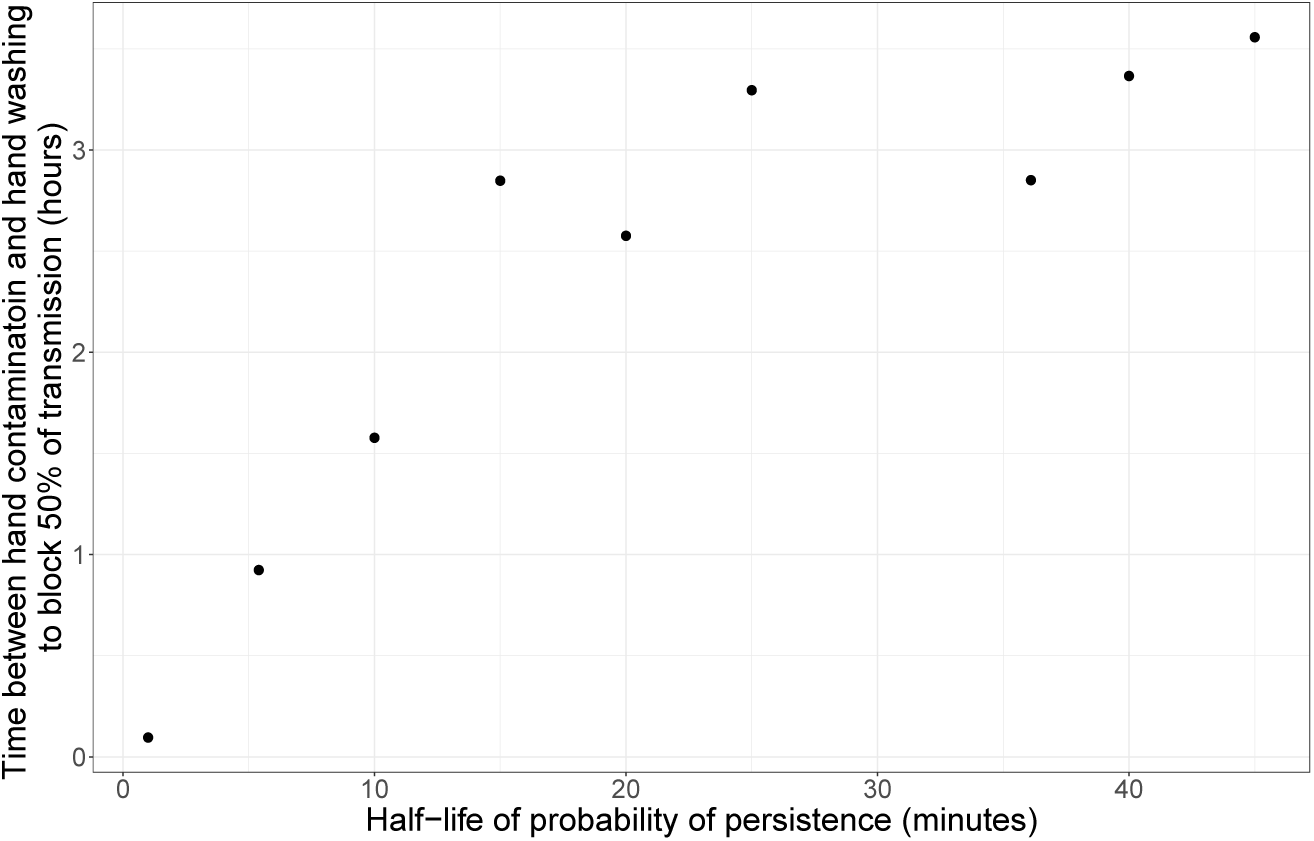
Cumulative time between hand contamination events and fixed-time hand washes to prevent 50% of transmissions. For each half-life value of virus persistence, the cumulative time between hand contamination events and hand washes for preventing 50% of transmission was computed for a baseline probability of infection of 10%. Hand contamination events are assumed to occur on average 4 times per hour.

## Transmission probability per contact and half-life of virus persistence

Figure S4 shows that the shorter the virus persists on hands, the higher the probability of transmission per face-touching contact has to be if the cumulative probability of infection is assumed to be fixed.

**Figure S4.**
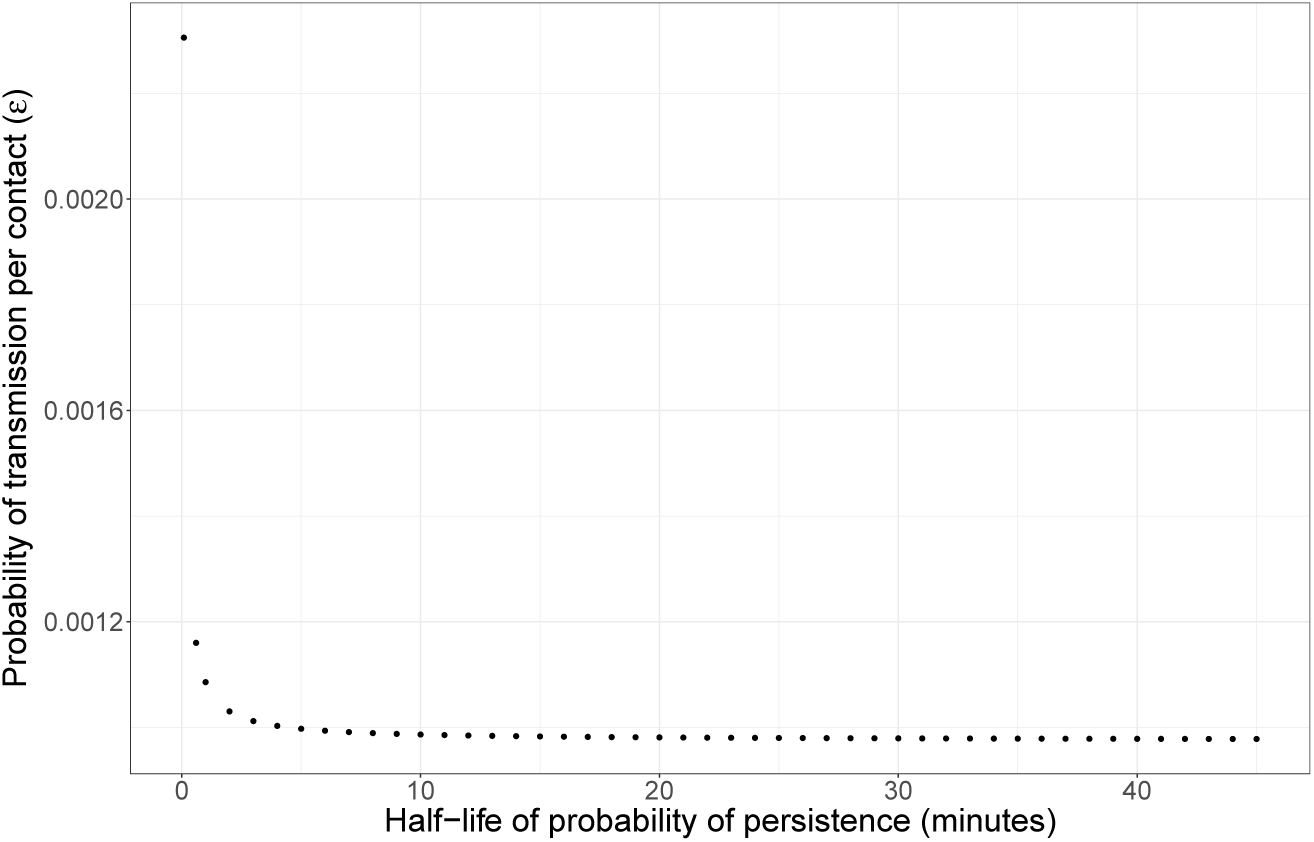
Probability of transmission per face-touching contact for different half-lives of virus persistence. For a baseline cumulative probability of infection of 10% and each half-life value of virus persistence, the probability of transmission per single face-touching contact was cmputed. Hand contamination events are assumed to occur on average 4 times per hour.

## Sensitivity analyses

### Cumulative probability of infection

We performed sensitivity analyses for different probabilities of infection and present here the results for probability of infection = 30% and probability of infection = 50% (see Figure S5-S6).

Figure S7 shows the impact of hand contamination rate on the number of hand washes that are necessary to prevent 50% of transmissions. A baseline probability of infection of 30% was used.

### Hand contamination event rate

We performed sensitivity analyses for different rates of hand contamination events and present the results for hand contamination rates of 1, 10 and 60 times per hour. The less frequently hands get contaminated, the larger the impact of increasing hand washing frequencies or reducing the delay of hand washing after hand contamination events and the larger the impact of the half-life of the probability of persistence of the virus on the actual probability of infection reduction. Figure S8 shows the results for a hand contamination rate of *λ_c_* =1 hour^-1^. The conclusions drawn from the Results section are applicable in this scenario as well. Figure S9-S10 depict the results for a hand contamination rate of 10 and 60 times per hour, respectively. When hand contamination occurs very frequently, fixed-time and event-prompted hand washing have almost identical effects. For both hand washing schemes, increasing the hand washing uptake has only a small impact on the probability of infection unless hand washing is performed every 5 minutes or the time delay of hand washing after hand contamination events is decreased to one or five minutes. However, due to the the high rate of hand contamination events of every 4 minutes or every minute, respectively, such an uptake seems infeasible. Hence, when susceptible individuals are exposed to continuous contamination, the best strategy would be to wash their hands as frequently as possible, especially after touching potentially contaminated surfaces, and to reduce the rate of contamination by, e.g., cleaning surfaces in their environment or isolating the infectious person.

**Figure S5.**
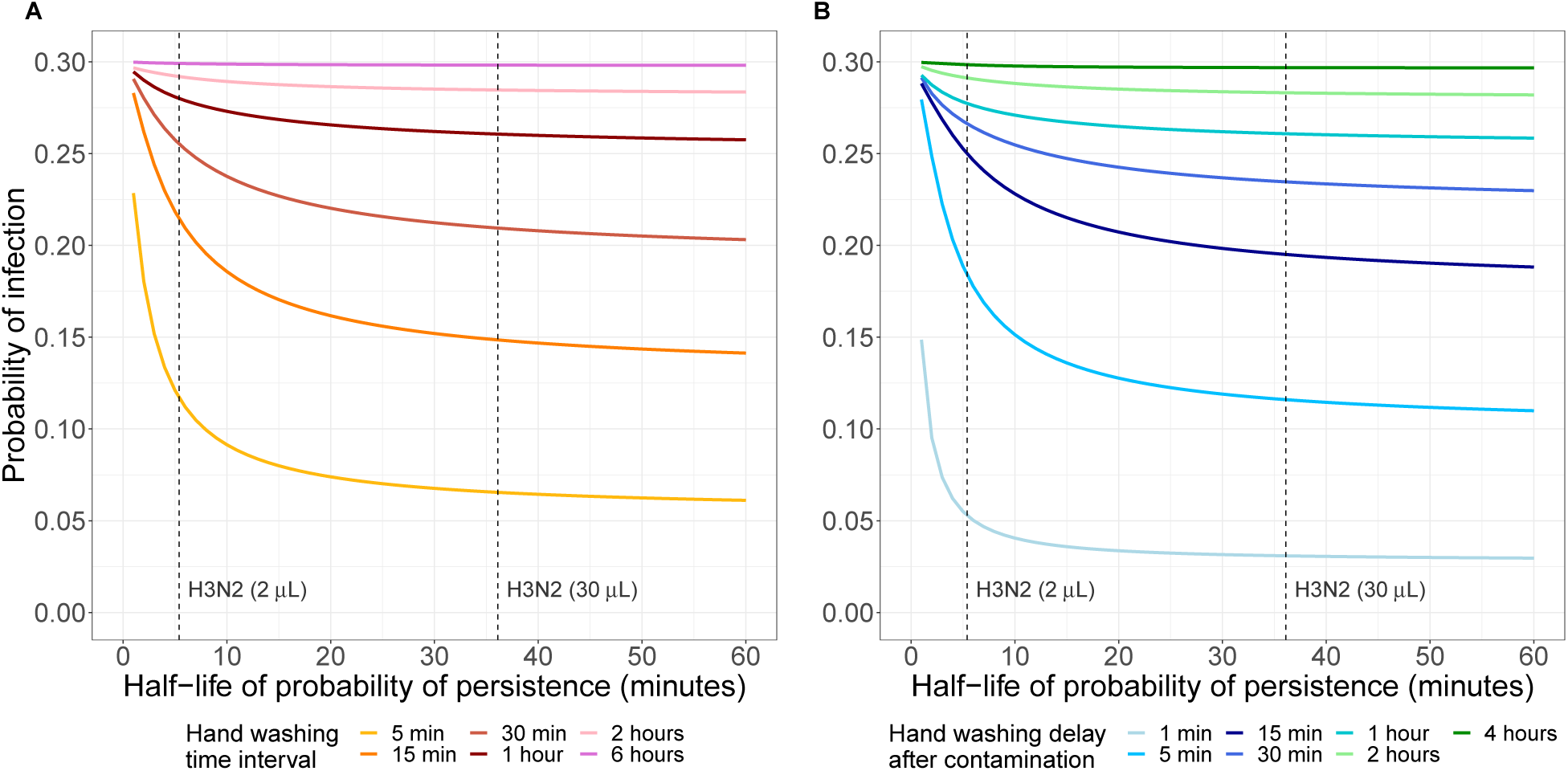
Impact of half-life of probability of virus persistence on probability of infection for different hand washing schemes and frequencies. (A) Fixed-time hand washing (B) Event-prompted hand washing. The dashed lines represent the half-life of probability of persistence for H3N2 for viral inoculum volumes of 2 *μ*L and 30 *μ*L (calculated from [19]). For each half-life value of the virus, the probability of transmission per face-touching event *ε* was determined for a probability of infection = 30% in the case of no hand washing. The probability of infection for the different hand washing frequencies/delays was then computed using this *ε* value. Hand contamination events are assumed to occur on average 4 times per hour.

**Figure S6.**
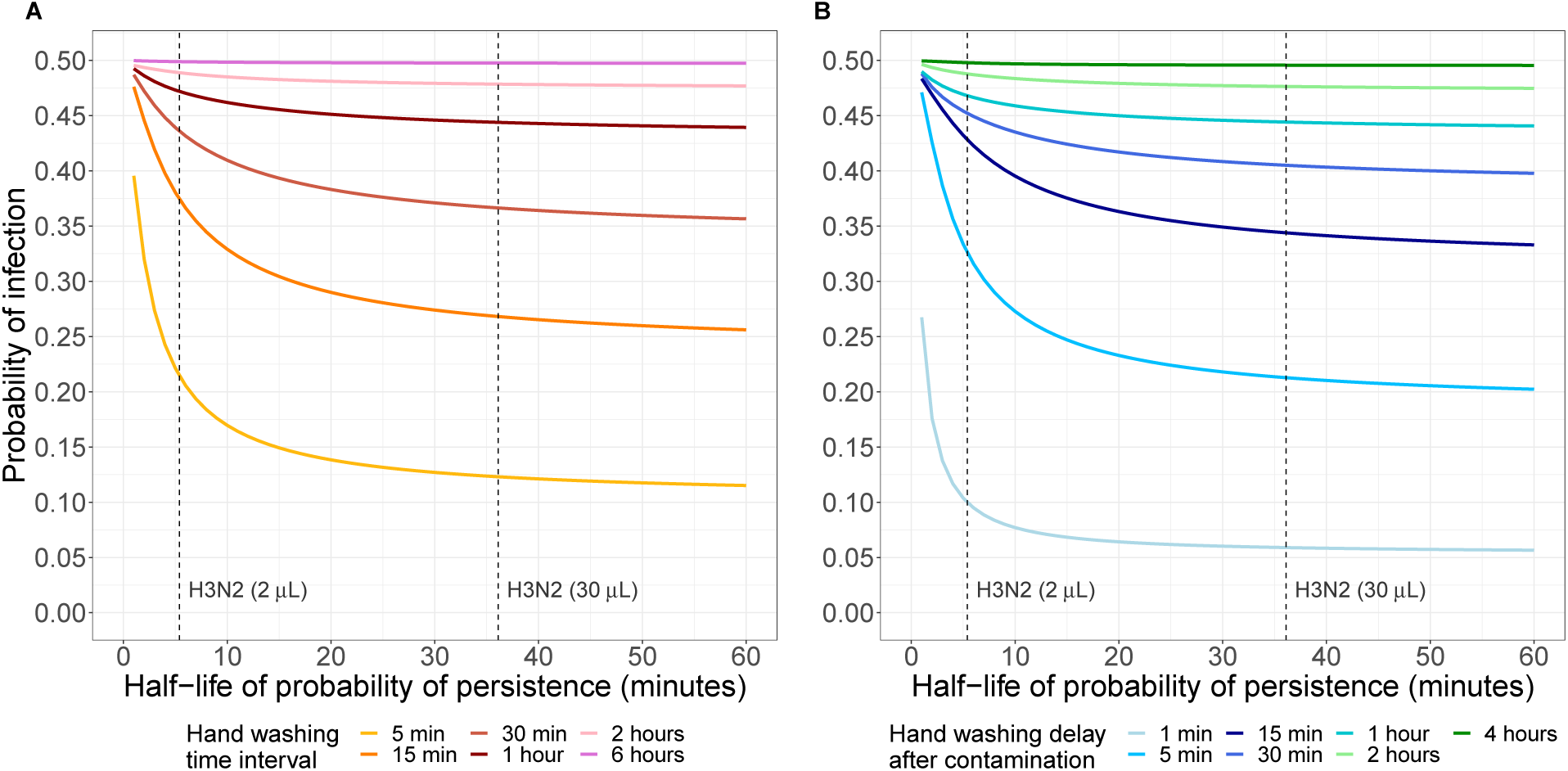
Impact of half-life of probability of virus persistence on probability of infection for different hand washing schemes and frequencies. (A) Fixed-time hand washing (B) Event-prompted hand washing. The dashed lines represent the half-life of probability of persistence for H3N2 for viral inoculum volumes of 2 *μ*L and 30 *μ*L (calculated from [19]). For each half-life value of the virus, the probability of transmission per face-touching event *ε* was determined for a probability of infection = 50% in the case of no hand washing. The probability of infection for the different hand washing frequencies/delays was then computed using this *ε* value. Hand contamination events are assumed to occur on average 4 times per hour.

**Figure S7.**
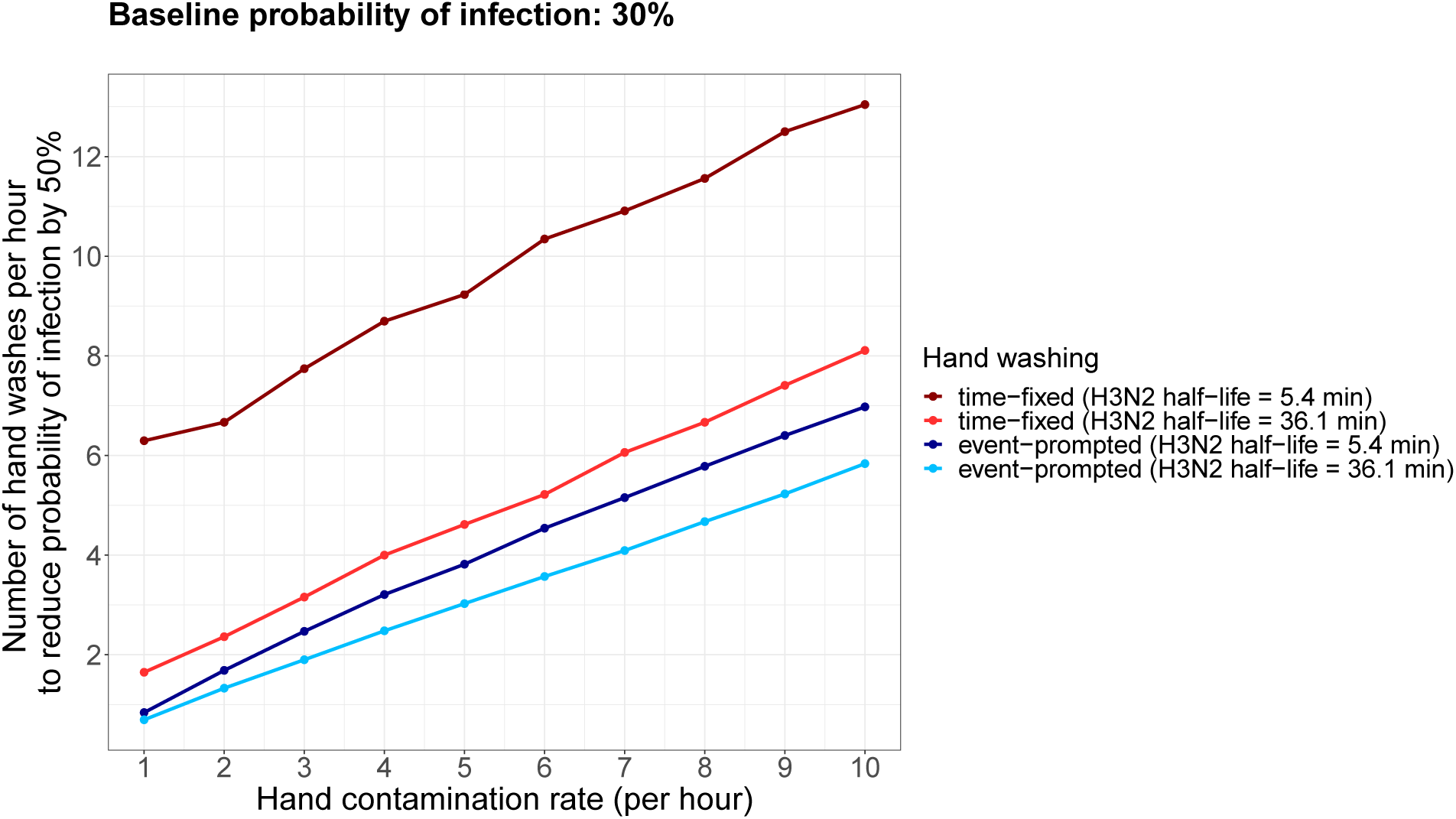
Number of hand washes necessary to decrease the cumulative probability of infection by 50%.

**Figure S8.**
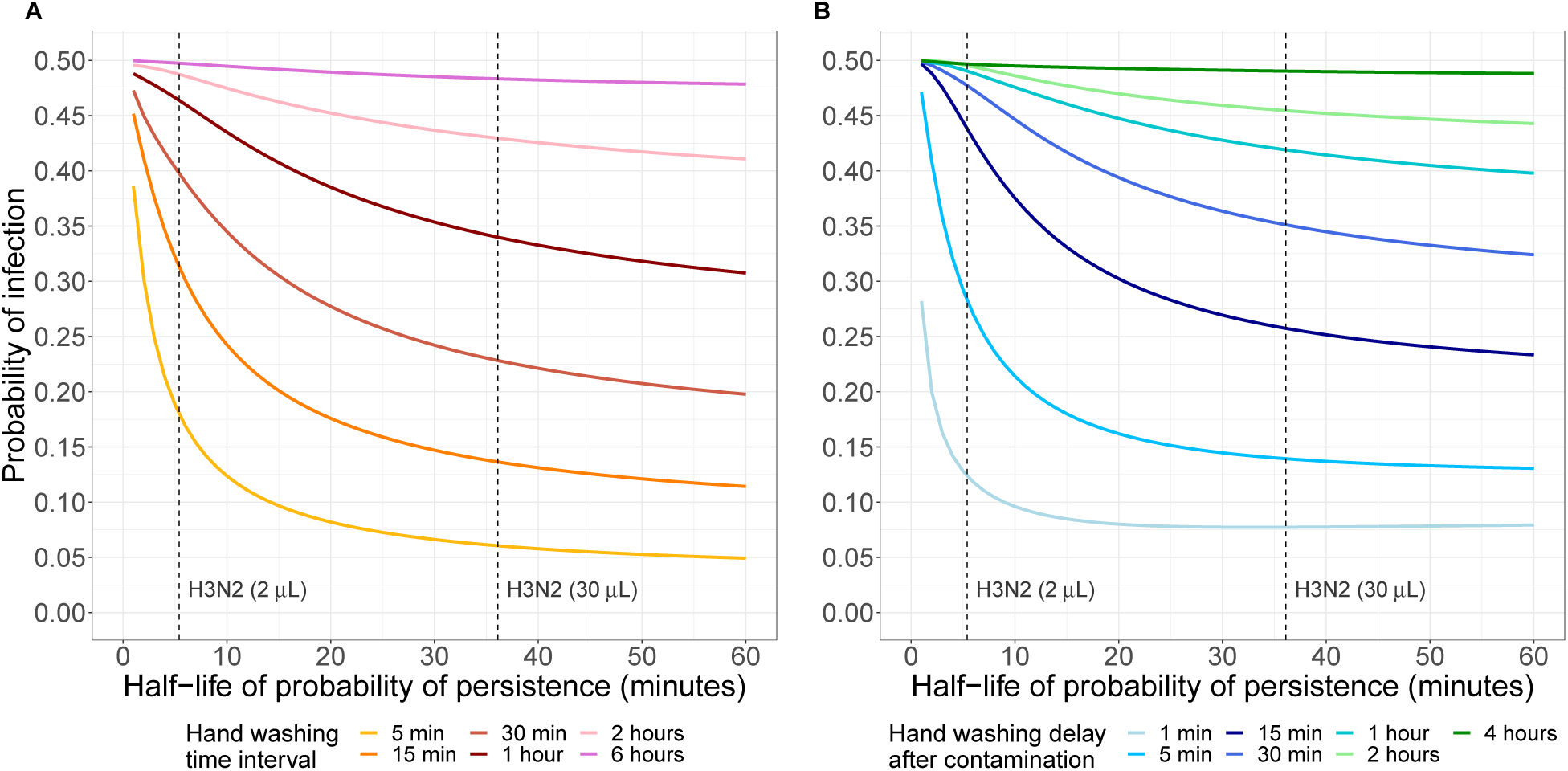
Impact of half-life of probability of virus persistence on probability of infection for different hand washing schemes and frequencies. (A) Fixed-time hand washing (B) Event-prompted hand washing. The dashed lines represent the half-life of probability of persistence for H3N2 for viral inoculum volumes of 2 *μ*L and 30 *μ*L (calculated from [19]). For each half-life value of the virus, the probability of transmission per face-touching event *ε* was determined for a probability of infection = 30% in the case of no hand washing. The probability of infection for the different hand washing frequencies/delays was then computed using this *ε* value. Hand contamination events are assumed to occur on average once per hour.

**Figure S9.**
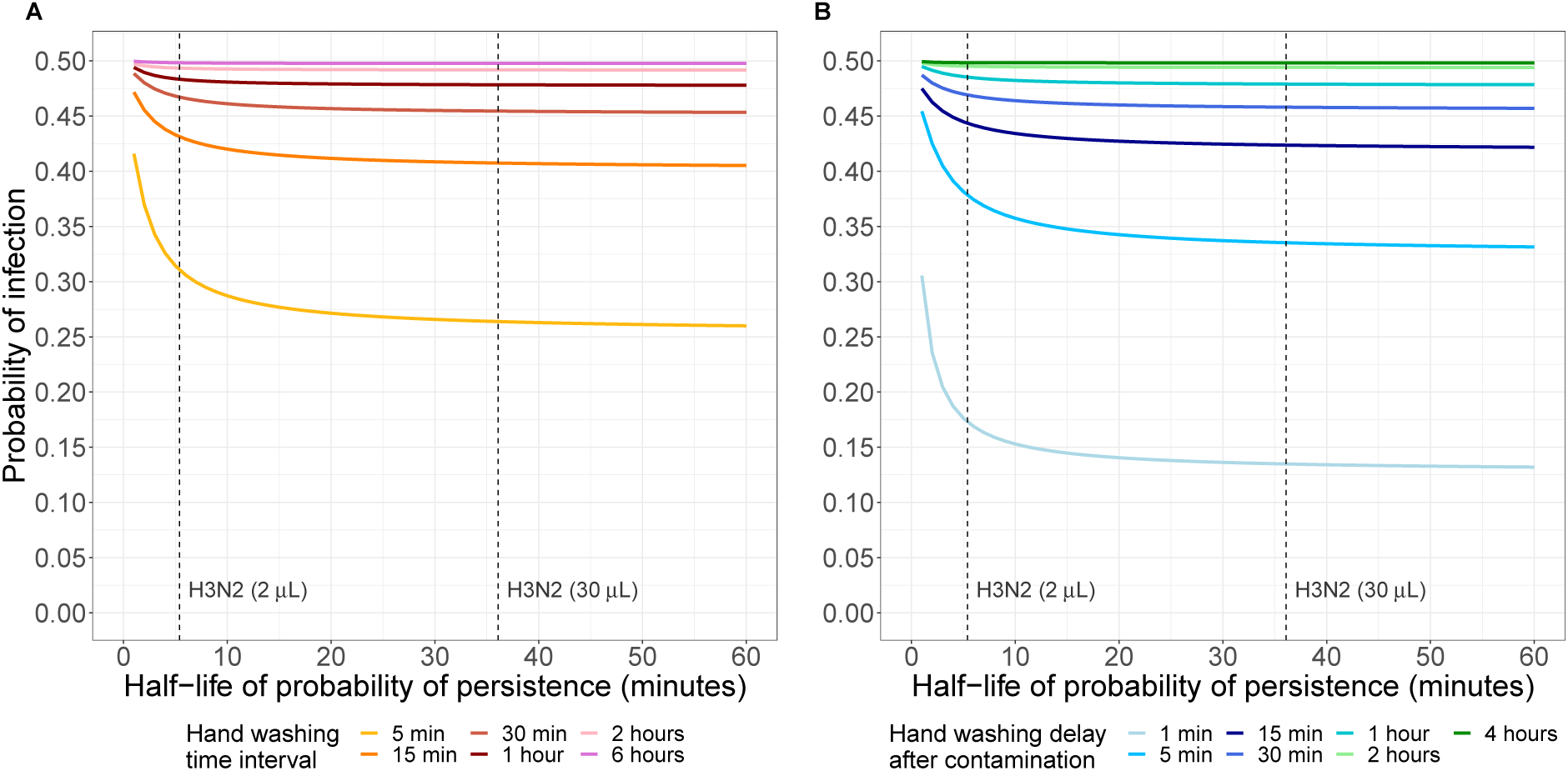
Impact of half-life of probability of virus persistence on probability of infection for different hand washing schemes and frequencies. (A) Fixed-time hand washing (B) Event-prompted hand washing. The dashed lines represent the half-life of probability of persistence for H3N2 for viral inoculum volumes of 2 *μ*L and 30 *μ*L (calculated from [19]). For each half-life value of the virus, the probability of transmission per face-touching event *ε* was determined for a probability of infection = 50% in the case of no hand washing. The probability of infection for the different hand washing frequencies/delays was then computed using this *ε* value. Hand contamination events are assumed to occur on average 10 times per hour.

### Comparison of number of hand washes

Figure S11 shows the average number of hand washes per hour for the two hand washing schemes in the scenario used in the main analysis, i.e. for a hand contamination rate *λ_c_* = 4 hour^-1^. For a fair comparison between the two hand washing schemes, fixed-time hand washing should be compared to event-prompted hand washing using approximately the same average number of hand washer per hour. For example, hand washing every fifteen minutes may be compared to event-prompted hand washing one minute after hand contamination.

**Figure S10.**
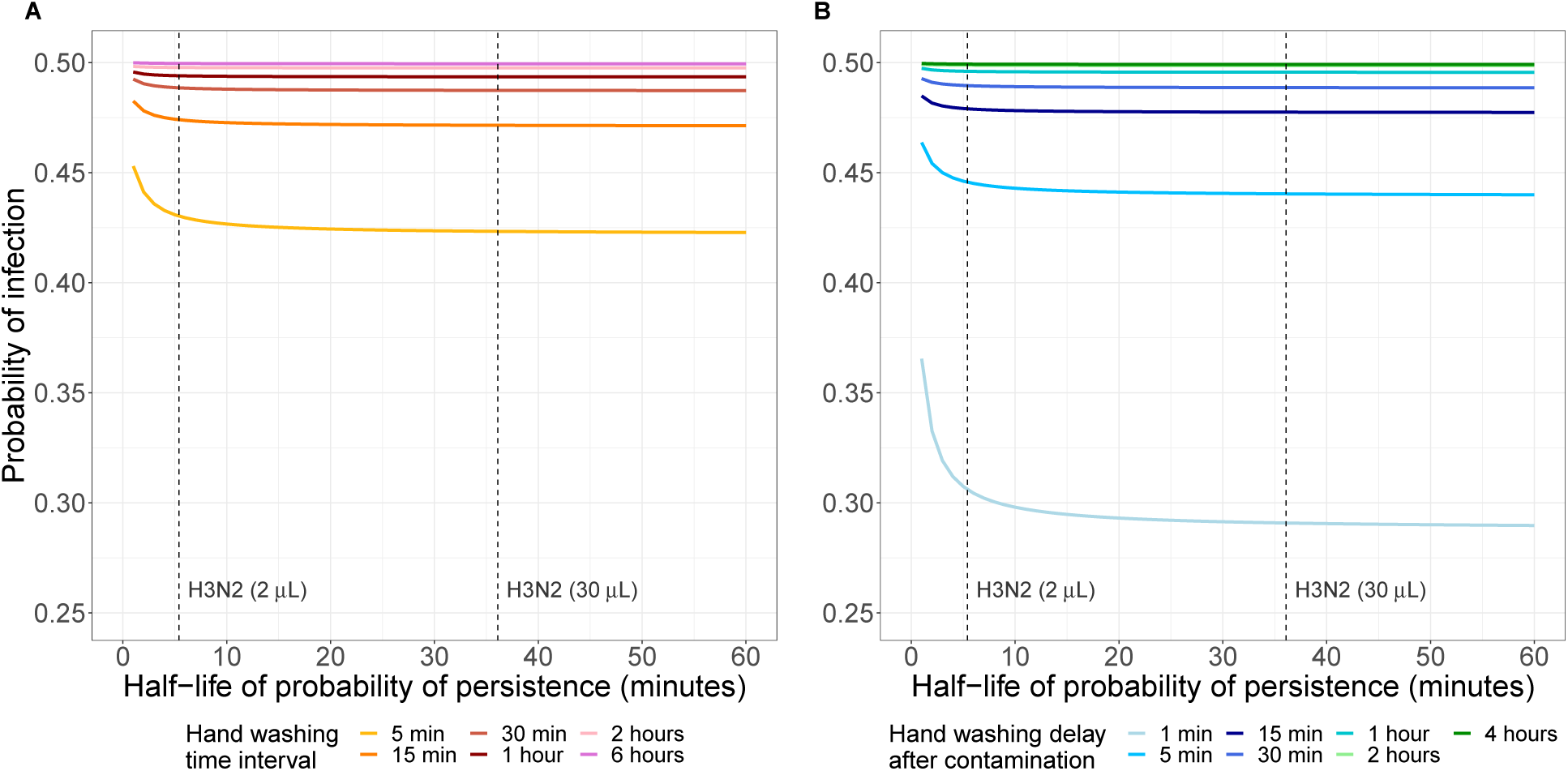
Impact of half-life of probability of virus persistence on probability of infection for different hand washing schemes and frequencies. (A) Fixed-time hand washing (B) Event-prompted hand washing. The dashed lines represent the half-life of probability of persistence for H3N2 for viral inoculum volumes of 2 *μ*L and 30 *μ*L (calculated from [19]). For each half-life value of the virus, the probability of transmission per face-touching event *ε* was determined for a probability of infection = 50% in the case of no hand washing. The probability of infection for the different hand washing frequencies/delays was then computed using this *ε* value. Hand contamination events are assumed to occur on average 60 times per hour.

**Figure S11.**
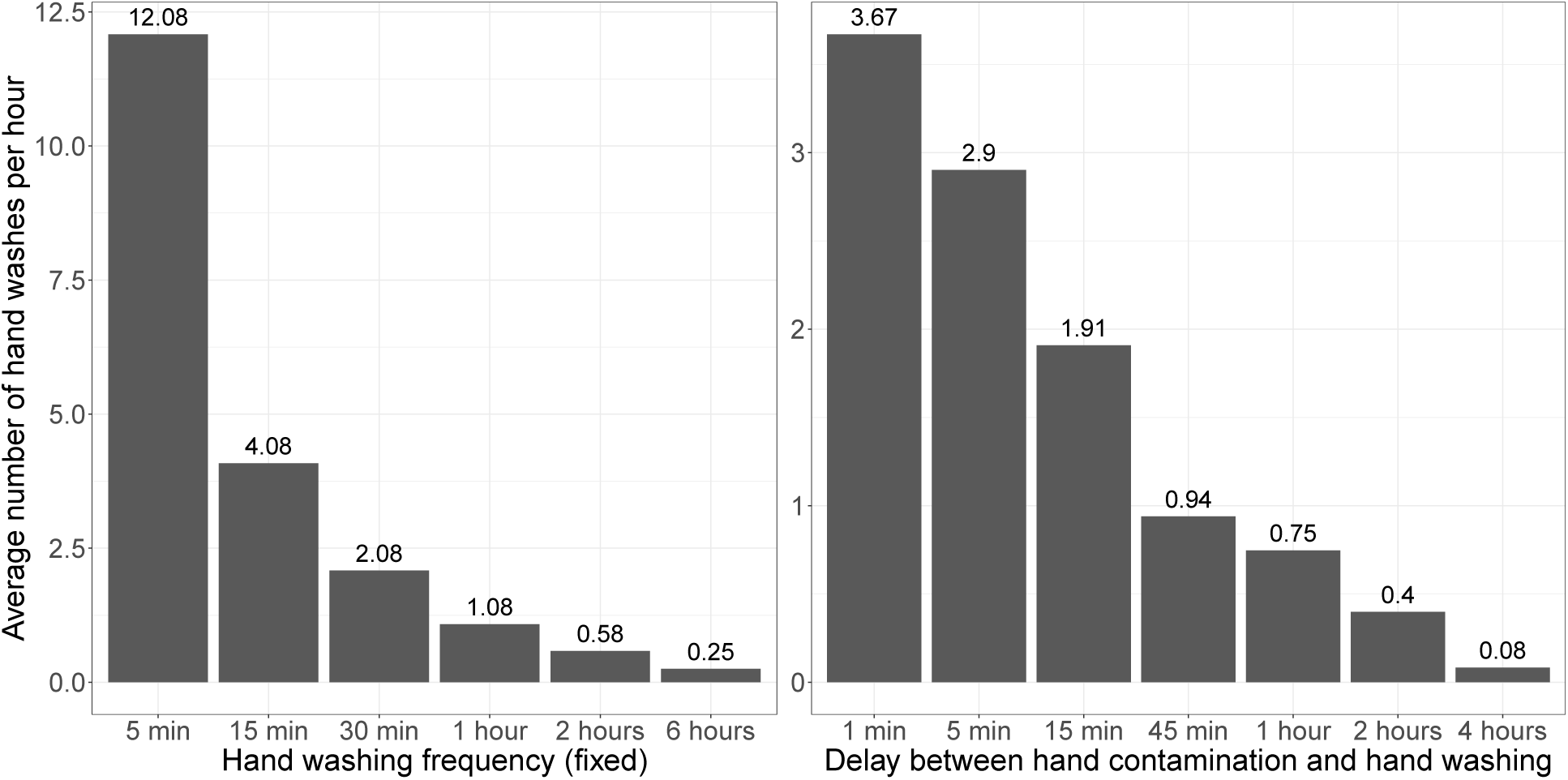
Average number of hand washes per hour for fixed and delayed hand washing. Hand contamination events are assumed to occur on average 4 times per hour.

## Notes

### Competing Interest Statement

The authors have declared no competing interest.

### Funding Statement

MY is supported by the Singapore National Medical Research Council Research Fellowship
311 (Grant ref: NMRC/Fellowship/0051/2017). TMP is supported by the Society for Laboratory
312 Automation and Screening (Grant ref: SLAS_VS2020). BSC is supported by the UK Medical
313 Research Council / Department for International Development (Grant ref: MR/K006924/1) and 314 the UK Department of Health and Social Care using UK Aid funding managed by the NIHR 315 (Grant ref: PR-OD-1017-20006). CL is supported by the Wellcome Trust (Grant ref: 316 206736/Z/17/Z). The funders had no role in study design, data collection and analysis, decision 317 to publish, or preparation of the manuscript. The views expressed in this publication are those of
318 the authors and not necessarily those of the above funders.

